# Estimating the effectiveness of first dose of COVID-19 vaccine against mortality in England: a quasi-experimental study

**DOI:** 10.1101/2021.07.12.21260385

**Authors:** Charlotte Bermingham, Jasper Morgan, Daniel Ayoubkhani, Myer Glickman, Nazrul Islam, Aziz Sheikh, Jonathan Sterne, A. Sarah Walker, Vahé Nafilyan

## Abstract

**Background:** Estimating real-world vaccine effectiveness is vital to assess the impact of the vaccination programme on the pandemic and inform the ongoing policy response. However, estimating vaccine effectiveness using observational data is inherently challenging because of the non-randomised design and the potential for unmeasured confounding.

**Methods:** We used a Regression Discontinuity Design (RDD) to estimate vaccine effectiveness against COVID-19 mortality in England, exploiting the discontinuity in vaccination rates resulting from the UK’s age-based vaccination priority groups. We used the fact that people aged 80 or over were prioritised for the vaccine roll-out in the UK to compare the risk of COVID-19 and non-COVID-19 death in people aged 75–79 and 80–84.

**Findings:** The prioritisation of vaccination of people aged 80 or above led to a large discrepancy in vaccination rates in people 80–84 compared to those 75–79 at the beginning of the vaccination campaign. We found a corresponding difference in COVID-19 mortality, but not in non-COVID-19 mortality, suggesting that our approach appropriately addresses the issue of unmeasured confounding factors. Our results suggest that the first vaccine dose reduced the risk of COVID-19 death by 52.6% (95% Cl 26.6–84.2) in those aged 80.

**Interpretations:** Our results support existing evidence that a first dose of a COVID-19 vaccine has a strong protective effect against COVID-19 mortality in older adults. The RDD estimate of vaccine effectiveness is comparable to previously published studies using different methods, suggesting that unmeasured confounding factors are unlikely to substantially bias these studies.

**Funding:** Office for National Statistics.

**Research in Context:** *Evidence before this study:* We searched PubMed for studies reporting on the ‘real-world’ effectiveness of the COVID-19 vaccination on risk of death using terms such as “COVID-19”, “vaccine effectiveness”, “mortality” and “death”. The relevant published studies on this topic report vaccine effectiveness estimates against risk of death ranging from 64.2% to 98.7%, for varying times post-vaccination. All of these are observational studies and therefore potentially subject to bias from unmeasured confounding. We found no studies that used a quasi-experimental method such as regression discontinuity design, which is not subject to bias from unmeasured confounding, to calculate the effectiveness of the COVID-19 vaccination on risk of COVID-19 death, or on other outcomes such as hospitalisation or infection.

*Added value of this study:* The estimates of vaccine effectiveness based on observational data may be biased by unmeasured confounding. This study uses a regression discontinuity design to estimate vaccine effectiveness, exploiting the fact that the vaccination campaign in the UK was rolled out following age-based priority groups. This enables the calculation of an unbiased estimate of the effectiveness of the COVID-19 vaccine against risk of death. The vaccine effectiveness estimate of 52.6% (95% Cl 26.6–84.2) is slightly lower but similar to previously published estimates, therefore suggesting that these estimates are not substantially affected by unmeasured confounding factors and confirming the effectiveness of the COVID-19 vaccine against risk of COVID-19 death.

*Implications of all the available evidence:* Obtaining an unbiased estimate of COVID-19 vaccine effectiveness is of vital importance in informing policy for lifting COVID-19 related measures. The regression discontinuity design provides confidence that the existing estimates from observational studies are unlikely to be substantially biased by unmeasured confounding.

## Introduction

On 8 December 2020, the UK launched an ambitious vaccination programme to combat the coronavirus (COVID-19) pandemic. As of 8 June 2021, 77.3% of the adult population (aged 18 or over) have received the first dose of a vaccine against COVID-19.^1^

Clinical trials have shown high vaccine efficacy for all currently UK authorised and deployed vaccines. For the BNT162b2 mRNA vaccine (Pfizer-BioNTech), 95% efficacy was reported against laboratory-confirmed COVID-19 [1]. The ChAdOx1 vaccine (Oxford-AstraZeneca) vaccine was found to have 70% efficacy against symptomatic COVID-19 amongst seronegative participants [2] The mRNA-1273 vaccine (Moderna), which was reported to have 95% efficacy against confirmed COVID-19, only started to be administered in the UK in April 2021 [3].

Monitoring real-world vaccine effectiveness is vital to assess the impact of the vaccination programme on the pandemic and inform the policy response. However, estimating real-world vaccine effectiveness without randomised control trials is challenging. Unvaccinated individuals are likely to differ from vaccinated individuals in ways that are not easy to measure, particularly when uptake is very high. Several studies have estimated vaccine effectiveness against the risk of infection, hospitalisation and death by comparing outcomes of vaccinated and unvaccinated individuals, adjusting for a range of individual characteristics. [4] [5] [6] [7] However, unmeasured confounders and temporal changes in the background infection rates may bias the estimates of vaccine effectiveness, towards overstating the effect. Indeed, many of these studies report large effectiveness in the first few days after vaccination, for instance because those who recently tested positive or self-isolating were asked to delay their vaccination.

If vaccine eligibility is based on a continuous variable such as age, regression discontinuity design (RDD) can be used to obtain unbiased estimates of vaccine effectiveness, even in the presence of unmeasured confounding factors [8] [9]. RDD approaches have been previously used to demonstrate that the childhood Bacillus Calmette-Guérin (BCG) is not effective against COVID-19, demonstrating that the correlation between BCG vaccination coverage and reduced impact of COVID-19 in different countries was due to unmeasured confounders [10]. RDD has also been used to investigate the effectiveness of the Human papillomavirus (HPV) vaccine against cervical dysplasia based on birth date [11]. In these examples, a policy change generated a cut-off in vaccine eligibility for a particular birth date; therefore, outcomes such as mortality could be compared around this cut-off to determine their relationship to vaccination. RDD has been proposed as a method to estimate COVID-19 vaccine effectiveness due to the age-based roll-out that many countries are adopting but has not hitherto been used [12].

In this study, we used an RDD to estimate vaccine effectiveness against COVID-19 mortality in England, exploiting the fact that the vaccination campaign in the UK was rolled out following age-based priority groups [13]. People aged 80 or over and health and social care professionals were targeted first by the vaccination campaign. Using data from the period where there was a substantial difference in the vaccination coverage of those aged just over 80 years compared to those just under 80 years, we used a fuzzy RDD to estimate the effect of vaccination on the risk of COVID-19 death. We also estimated the effect of vaccination on the risk of non-COVID-19 death as a falsification test. We then calculate vaccine effectiveness based on the results from the fuzzy RDD.

## Methods

### Study data

We linked vaccination data from the National Immunisation Management System (NIMS) to the Office for National Statistics (ONS) Public Health Data Asset (PHDA) based on NHS number. The ONS PHDA is a linked dataset combining the 2011 Census, mortality records, the General Practice Extraction Service (GPES) data for pandemic planning and research and the Hospital Episode Statistics (HES). To obtain NHS numbers for the 2011 Census, we linked the 2011 Census to the 2011-2013 NHS Patient Registers using deterministic and probabilistic matching, with an overall linkage rate of 94.6%. All subsequent linkages were performed based on NHS numbers.

The study population consisted of people aged 75–84 years, alive on 8 December 2020, who were resident in England, registered with a general practitioner, and enumerated at the 2011 Census. Of 3,322,785 adults aged 75–84 years who received a first dose of a COVID-19 vaccine in NIMS by 3 May 2021, 3,118,492 (93.9%) were linked to the ONS PHDA.

### Outcomes

Vaccine effectiveness was estimated against mortality involving COVID-19. A death was defined as involving COVID-19 if COVID-19 was recorded as an underlying or contributory cause of death on the death certificate as identified by either ICD-10 (International Classification of Diseases, 10th revision) codes U07.1 (confirmed) or U07.2 (suspected). Mortality where COVID-19 was not mentioned on the death certificate was also investigated as falsification test, to provide a check for bias in the results.

### Intervention

Treatment was defined as having received a first dose of a vaccine against COVID-19 (Pfizer-BioNTech or Oxford-AstraZeneca) for long enough to be likely to have reached seropositivity and be protected. The expected proportion of people reaching seropositivity at different lengths of time post-vaccination was estimated from a study investigating antibody response following vaccination using data from the COVID-19 Infection Survey [14]. In the survey, SARS-CoV-2 antibody levels are measured using an ELISA detecting anti-trimeric spike IgG using a fluorescence detection mechanism to 26 February 2021, then a CE-marked version of the assay, the Thermo Fisher OmniPATH 384 Combi SARS-CoV-2 IgG ELISA, using the same antigen with a colorimetric detection system. A threshold of ≥42 ng/ml was used to identify IgG-positive samples. We used these estimates of percentages reaching seropositivity by age and by time from vaccination to define the treatment indicator.

We calculated the proportion of people likely to reach the threshold level for seropositivity in our dataset by constructing cumulative incidence curves for vaccination a certain number of days prior and adjusting by the proportion of people expected to reach seropositivity for that number of days post-vaccination using the COVID-19 Infection Survey data for people aged 80 [7]. Pfizer-BioNTech and Oxford-AstraZeneca vaccinations were weighted separately to account for the differing response curves of these vaccines. The percentages of 80 year olds reaching the threshold antibody level 28 days after the first vaccination dose were 85.2% (95% Cl 80.5–88.9) and 73.7% (95% Cl 65.9–80.3) for those vaccinated with the Pfizer-BioNTech and Oxford-AstraZeneca vaccinations respectively. A weighted value for the proportion treated for each age in months group was calculated over the analysis period, accounting for the time elapsed since vaccination and the changes over the analysis period. More details are presented in the Supplementary Appendix 1.

We used this threshold antibody level to define treatment, rather than vaccination, because protection is likely to be limited until this level was reached. Also of relevance is that for the RDD, we were interested in the overall level of protection that the vaccine provides in the population.

### Eligibility

The vaccination programme in the UK was rolled out following age-based priority groups, with people aged 80 or over being targeted first [13]. Therefore, eligibility for the vaccine was defined as being aged 80 or over at the beginning of the vaccination campaign (8 December 2020).

### Statistical analyses

We aimed to estimate the effect on the risk of COVID-19 mortality of being protected by first dose of a vaccine (defined as being vaccinated for long enough to be likely to have reached seropositivity) for people aged 80. To do so, we used a fuzzy RDD, exploiting the fact that people aged 80 or over were prioritised for vaccination at the start of the vaccination campaign, but the compliance was imperfect [15]. Because those aged 75–79 rapidly became eligible for vaccination, we restricted the analysis window to the period where there was a large difference in vaccination rates between those above and below 80 years old.

We estimated the discontinuities in treatment and outcome over 15-day periods to ensure sufficient data to estimate the effect of being protected on COVID-19 mortality, with start dates ranging from 7 January December 2020 to 26 February 2021. We started the analytical window 30 days after the start of the vaccination campaign, allowing time for vaccinated people to become protected. By the end of February 2021, the difference in vaccination rates between peopled aged 75–79 and 80–84 was very small. For our main analysis, we selected the period where the probability of being vaccinated and having reached antibody threshold for those aged above 80 years was approximately five times of that for those aged under 80 years (16 to 30 January 2021). We also reported results for alternative analysis periods.

First, we estimated the effect of eligibility (being aged 80 or over at the beginning of the roll out of the vaccination programme) on the probability of being protected. We calculated the proportion of people who were protected, by age in months. We then estimated the discontinuity in vaccination at the cut-off by fitting a linear regression model, adjusted for age in month, interacted with a binary variable for being aged 80 over, to allow for the effect of age to vary on both sides of the cut-off, as is standard practice in RDD studies [15].

Second, we calculated the effect of eligibility (being aged 80 or over at the beginning of the roll out of the vaccination programme) on the 15-day COVID-19 mortality rates by age in month groups, by deriving multi-state cumulative incidence curves for deaths involving COVID-19 and for deaths not involving COVID-19 for all people in our sample alive on the first day of the analysis period (16 January). We then estimated the discontinuity in the 15-day COVID-19 mortality rate at the cut-off by fitting a linear regression model, adjusted for age in month, interacted with a binary variable for being aged 80 or over.

Finally, using the proportion of people who were protected, by age in months we estimated the effect of vaccination on COVID-19 mortality by fitting a fuzzy RDD, using eligibility (being 80 or over) as an instrumental variable for the treatment. We thereby obtained an estimate of the Local Average Treatment Effect (LATE).

To calculate the vaccine effectiveness, we fitted the same model but used the log odds of COVID-19 mortality as the dependent variable instead of the COVID-19 mortality rate. This allows us to estimate the odds ratio (OR) of being vaccinated, and to derive the vaccine effectiveness as one minus the OR. All analyses were weighted by the number of people in that age group in the analysis dataset.

### Sensitivity analyses

The validity of the RDD rests on the assumption that in the absence of the intervention there would have been no discontinuity in the outcome. Whilst this assumption cannot be tested directly, we checked for continuity across the eligibility cut-off in pre-determined characteristics, such as sex, quintiles of Index of Multiple Deprivation (IMD) and prevalence of a comorbidity (Supplementary Appendix 1).

In our main analysis, we estimate the LATE of vaccination on mortality using data for people aged 75–84, hence using a bandwidth of five years (on each side of the eligibility cut-off). As a sensitivity analysis, we estimate the LATE using different bandwidths, from 10–60 months. (Supplementary Appendix 2). A smaller bandwidth may reduce bias, but may result in less precise estimates. We also removed people very close to the cut-off, as their eligibility status could be misclassified (Supplementary Appendix 3).

To check that the discontinuity in COVID-19 mortality rates is caused by eligibility to the vaccine, we estimated discontinuity in outcomes at arbitrarily chosen cut-offs, where we would expect no discontinuity (Supplementary Appendix 4). We also present results for different analysis periods (In Results section and Supplementary Appendix 5).

## Results

Characteristics of the study population are shown in Table 1. Out of the 3,422,644 people aged 75–84 included in our study, 91.2% had received a first dose of a COVID-19 vaccine by 3 May 2021. 54.3% were women; the average age at 8 December 2020 was 79.0 and 44.9% had at least one long-term health problem known to increase the risk of severe Covid outcomes (as defined as per the QCovid risk model) [16].

**Table 1:**
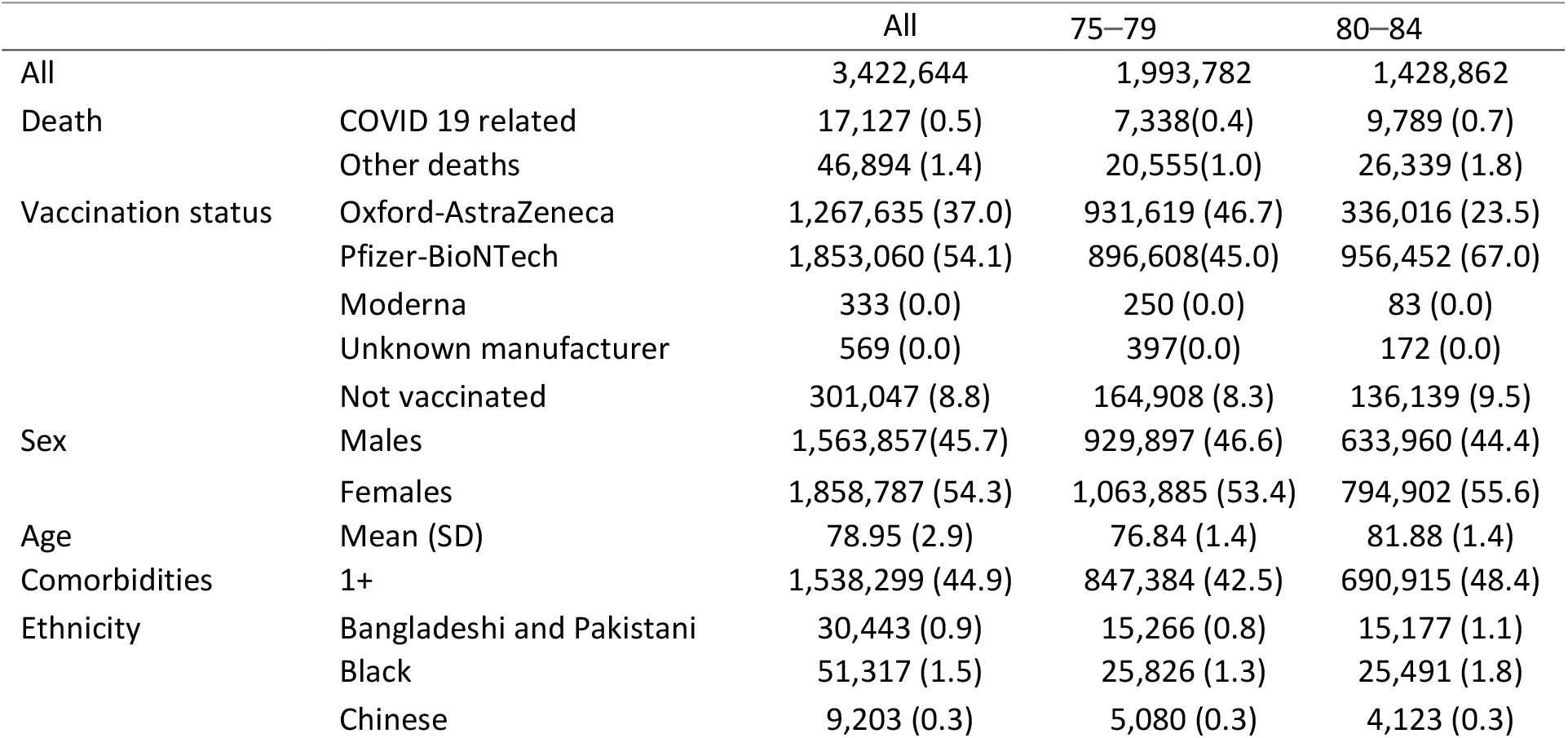
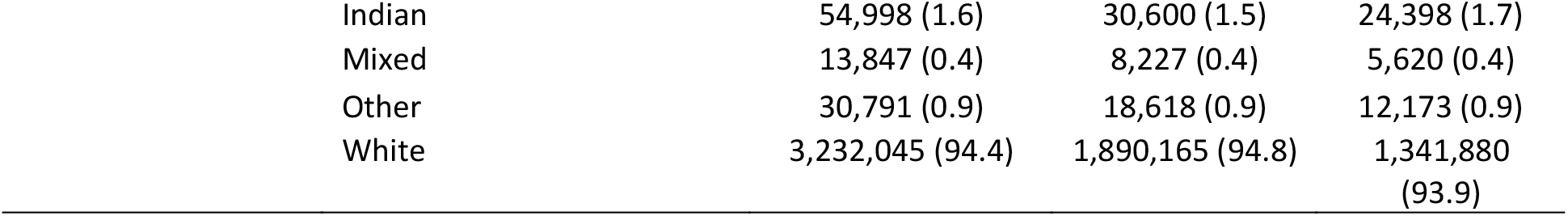
Characteristics of study population.

|  |  | All | 75–79 | 80–84 |
| --- | --- | --- | --- | --- |
| All |  | 3,422,644 | 1,993,782 | 1,428,862 |
| Death | COVID 19 related | 17,127 (0.5) | 7,338(0.4) | 9,789 (0.7) |
|  | Other deaths | 46,894 (1.4) | 20,555(1.0) | 26,339 (1.8) |
| Vaccination status | Oxford-AstraZeneca | 1,267,635 (37.0) | 931,619 (46.7) | 336,016 (23.5) |
|  | Pfizer-BioNTech | 1,853,060 (54.1) | 896,608(45.0) | 956,452 (67.0) |
|  | Moderna | 333 (0.0) | 250 (0.0) | 83 (0.0) |
|  | Unknown manufacturer | 569 (0.0) | 397(0.0) | 172 (0.0) |
|  | Not vaccinated | 301,047 (8.8) | 164,908 (8.3) | 136,139 (9.5) |
| Sex | Males | 1,563,857(45.7) | 929,897 (46.6) | 633,960 (44.4) |
|  | Females | 1,858,787 (54.3) | 1,063,885 (53.4) | 794,902 (55.6) |
| Age | Mean (SD) | 78.95 (2.9) | 76.84 (1.4) | 81.88 (1.4) |
| Comorbidities | 1+ | 1,538,299 (44.9) | 847,384 (42.5) | 690,915 (48.4) |
| Ethnicity | Bangladeshi and Pakistani | 30,443 (0.9) | 15,266 (0.8) | 15,177 (1.1) |
|  | Black | 51,317 (1.5) | 25,826 (1.3) | 25,491 (1.8) |
|  | Chinese | 9,203 (0.3) | 5,080 (0.3) | 4,123 (0.3) |

|  |  |  |  |
| --- | --- | --- | --- |
| Indian | 54,998 (1.6) | 30,600 (1.5) | 24,398 (1.7) |
| Mixed | 13,847 (0.4) | 8,227 (0.4) | 5,620 (0.4) |
| Other | 30,791 (0.9) | 18,618 (0.9) | 12,173 (0.9) |
| White | 3,232,045 (94.4) | 1,890,165 (94.8) | 1,341,880 (93.9) |

Cumulative incidence curves for the proportion of people aged 75–79 years and 80–84 years who have received a first dose of a COVID-19 vaccine are shown in Figure 1. Due to the earlier administering of vaccinations for people aged 80+, there was a clear difference in the proportion of each age group that was vaccinated at the beginning of the vaccination campaign. On 8 January 2021, one month after the start of the vaccination roll out, 34% of people aged 80–84 had received a first dose of a vaccine, compared to 5% of those aged 75–79. The vaccination was then extended to younger age groups. On 8 February 2021, 92% of people aged 80–84 and 90% of those aged 75–79 had received their first dose; 13% of people aged 80–84 and 0.5% of those aged 75–79 had received their second dose (See Supplementary Figure S1). In addition, the rapid increase in the proportions vaccinated in both groups can be seen, which was taken into account when calculating the proportion protected for the analysis period.

**Figure 1.**
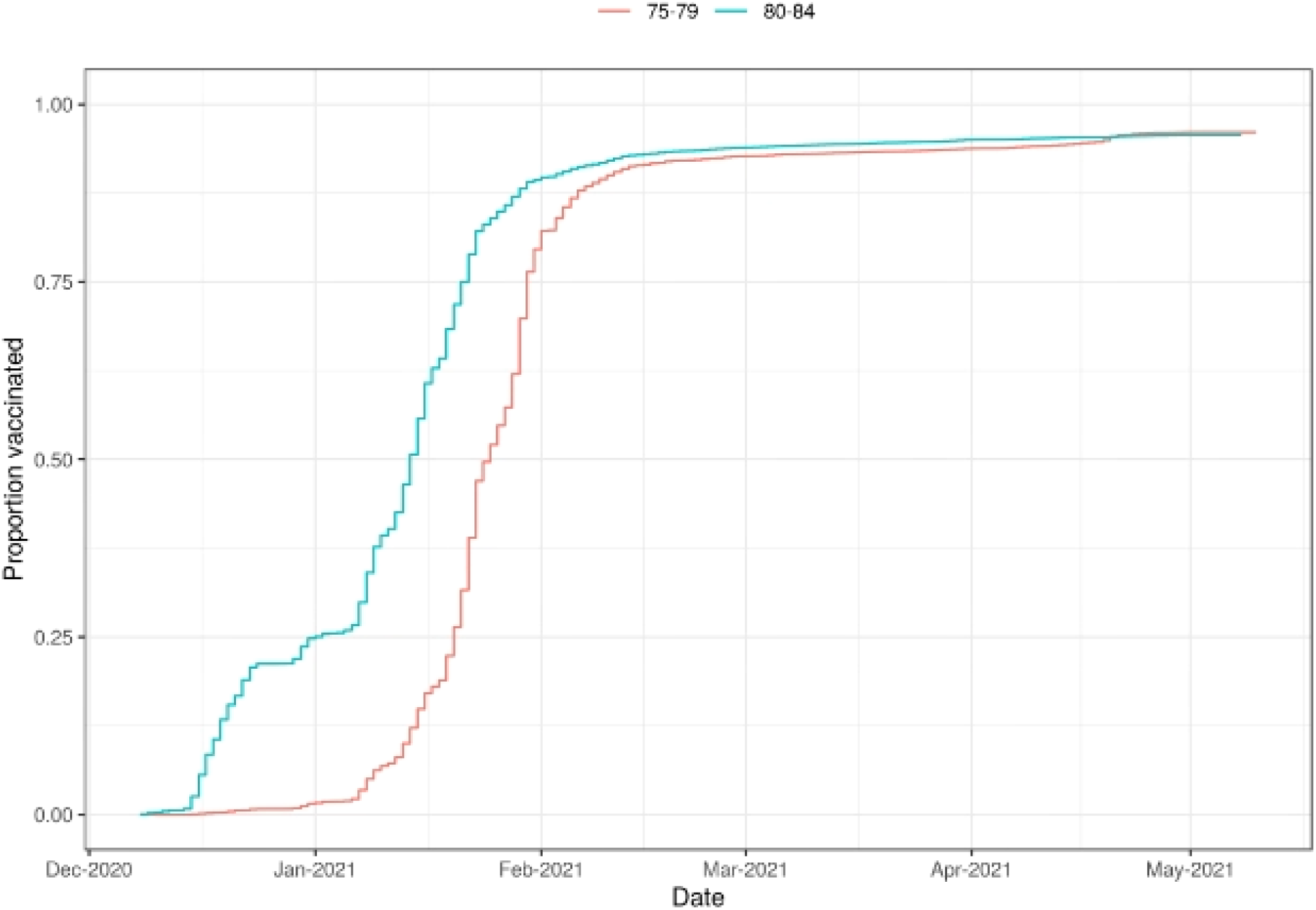
Cumulative incidence of first dose COVID-19 vaccination by age group (75*–*79, 80–84 years) from 8 December 2020 (date of first UK vaccination) to 3 May 2021. *Note: Cumulative incidence of first dose COVID-19 vaccination, with death treated as a competing risk. Cumulative incidence of second dose COVID-19 vaccination is reported in Supplementary Figure S1*.

### Main results

The weighted proportion protected (vaccinated and likely to have reached threshold antibody level) by age in month for the analysis period is shown in Panel A of Figure 2. The proportion of people having been vaccinated and reached threshold antibody level is much lower amongst people aged 75–79 than aged 80 or over, who had been prioritised for the vaccination. There is evidence of a clear discontinuity at age 80, with people near the eligibility cut-off being 5.5 times more likely to be protected just above than just below.

**Figure 2.**
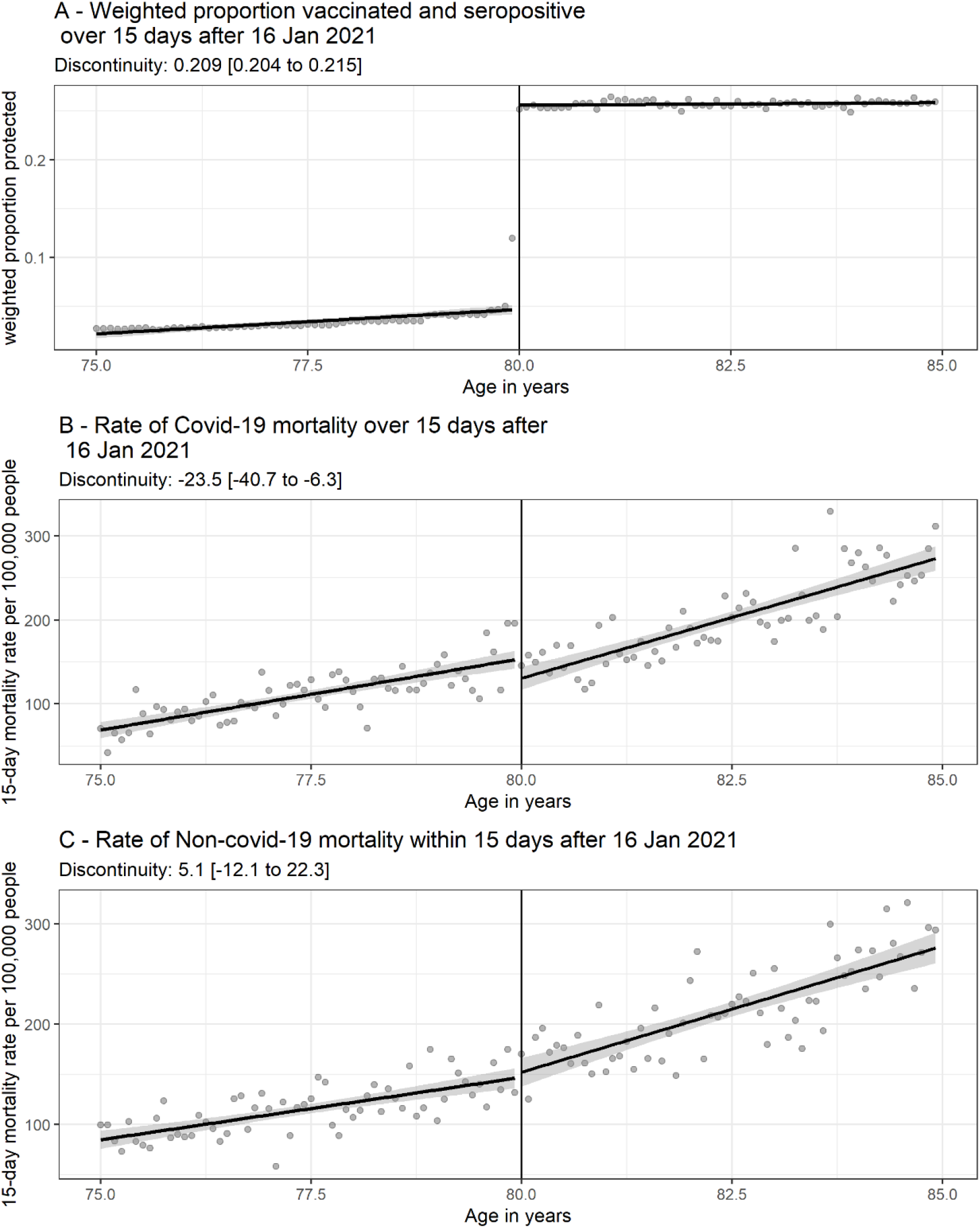
Regression discontinuity design plots for treatment and outcome variables by age in months for the 15-day period starting 16 Jan 2021. *Note:(A) Weighted proportion of people who have been vaccinated and reached threshold antibody level over the 15-day period by age in months. The discontinuity in the weighted proportion with antibodies at age 80 years is 0*.*209 (95% Cl 0*.*204–0*.*215, p value < 0*.*0001). (B) Probability of COVID-19 mortality during the 15-day period. The discontinuity in probability of COVID-19 mortality at age 80 years was -23*.*5 (95% Cl -40*.*7–6*.*3, p value 0*.*008) per 100,000 days at risk (C) Probability of non-COVID-19 mortality during the 15-day period. There is not a statistically significant discontinuity in the probability of non-COVID-19 mortality at age 80 years (p value 0*.*6). All data are fit using a linear regression model, adjusted for age in month, interacted with a binary variable for being aged 80 or over*.

The probability of COVID-19 mortality and mortality from other causes are shown in Panels B and C of Figure 2 respectively. A decrease in the probability of COVID-19 mortality can be seen at age 80. Being eligible for vaccination (e.g. being 80 or over) led to a reduction in the risk of COVID-19 mortality of 23.5 (95% CI 40.7–6.3) per 100,000 days at risk. As expected, there was no discontinuity in probability of mortality from other causes. This provides reassurance that the relationship between treatment and reduced COVID-19 mortality was not due to residual confounding.

The results from the fuzzy RDD, using eligibility as an instrument for being protected, indicate that receiving a first dose of a vaccine and reaching seropositivity reduced COVID-19 mortality by 112 (95% Cl 33–191) deaths per 100,000 people in the 15 days from 16 January 2021.

This yields a vaccine effectiveness of 52.6% (95% Cl 26.6–84.2) for people aged 80 who have been vaccinated for long enough to be likely to have reached seropositivity and therefore be protected.

The estimated discontinuity at age 80 in the weighted proportion of people who were protected for different analysis periods are reported in Figure 3A. The discontinuity at age 80 widened at the beginning of the vaccination campaign, as the 80+ group was prioritised. It then fell as the vaccination campaign was extended to people below 80. The effect of eligibility on the 15-day COVID-19 mortality rate (Figure 3B) also varied depending on the chosen analysis period due to the changing relative proportions of people protected in the eligible and ineligible groups and the underlying COVID-19 infection rate. The discontinuities in non-COVID-19 mortality were all not different from zero (Figure 3C). The estimates of the effect of receiving a first dose of a vaccine and reaching seropositivity on COVID-19 mortality for people aged 80, were relatively stable (Figure 3D) and varied with the underlying COVID-19 infection rate. The estimates of vaccine effectiveness are mostly consistent with the wide confidence intervals on the final estimate for the 16 January 2021 analysis (Figure 2F). The COVID-19 mortality rate in the unprotected population increased for later time periods. As more people became vaccinated, the unprotected population shrank and became different from the general population. The estimates therefore became less representative of the COVID-19 mortality rate in the general population for unvaccinated people for these analysis periods.

**Figure 3.**
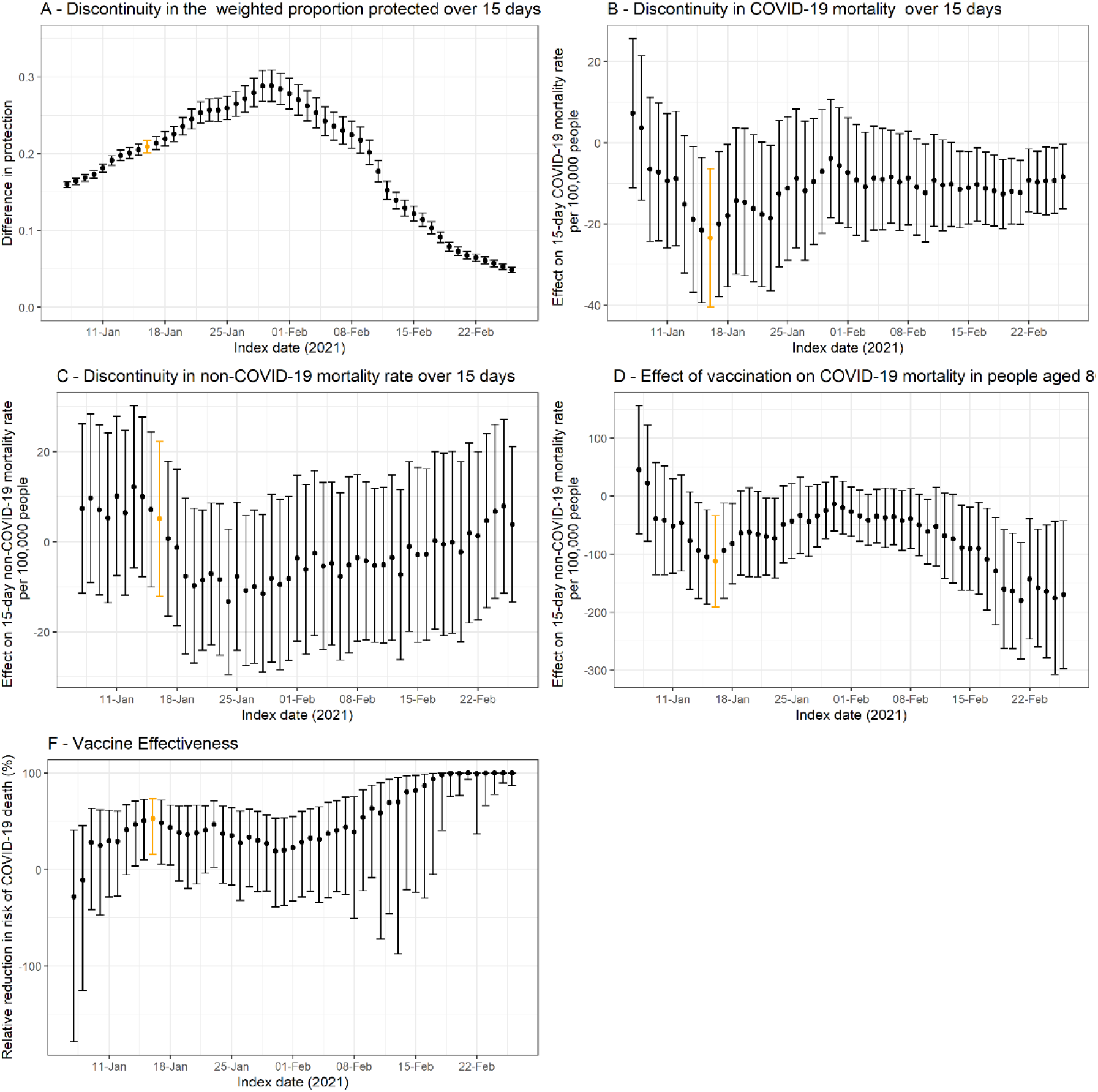
Estimated discontinuities, Local Average Treatment Effects and Vaccine effectiveness for alternative analysis periods. Note: (A) Discontinuity in the weighted proportion of people who have been vaccinated and reached threshold antibody level over the 15-day period (B) Discontinuity in the 15 day COVID-19 mortality rate (C) Discontinuity in the 15 day non-COVID-19 mortality rate (D) Effect of vaccination on COVID-19 mortality in people aged 80 (= (B) / (A)) (E) Vaccine effectiveness (= 1 – Odds ratio for being protected).

### Sensitivity analyses

Sensitivity analyses presented in the Supplementary Appendices demonstrated the applicability and robustness of RDD as a technique for estimating the vaccine effectiveness. Predetermined characteristics, such as sex, IMD quintile and prevalence of comorbidities, were continuous across the eligibility cut-off at 80 years (Supplementary Appendix 1). The discontinuity in the outcome variable was directly related to the discontinuity in the treatment variable and not influenced by confounding factors. We also found that the discontinuity in the outcome variable (COVID-19 mortality) occurred only at the eligibility cut-off of 80 years, and not at any other age in our age range, testing in 10-month steps from the eligibility cut-off, further confirming the relationship between the outcome and treatment discontinuities (Supplementary Appendix 4). There was no discontinuity in the probability of non-COVID-19 mortality at the eligibility cut-off nor at any other age cut-off.

The bandwidth used in our main analysis (60 months either side of the cut-off) resulted in the same value for the LATE as a bandwidth of 50 months, but smaller bandwidths led to less precisely estimated LATE (Supplementary Appendix 3). Excluding datapoints close to the cut-off did not significantly affect the results.

A longer period introduced more variation due to the changing vaccination status of the population and the changing infection levels. Shorter periods reduced the precision of the estimates because they did not contain enough mortality data to calculate reliable mortality estimates by age in months (Supplementary Appendix 5). A 15-day period reduces the relative uncertainty in the LATE compared to shorter periods. However, periods more than 15 days did not further reduce uncertainty, therefore indicating that there was sufficient mortality data to calculate the probabilities of COVID-19 mortality by age in months required to estimate the LATE.

## Discussion

In this study, we used an RDD approach to determine the effectiveness of the vaccination programme in England in preventing COVID-19 mortality for the population with age close to 80 years. By using a RDD approach, we obtained an estimate of vaccine effectiveness that was not biased by confounding factors, whether measured or not. Our results demonstrate a causal impact of vaccinations on reducing COVID-19 mortality, with effectiveness slightly lower but comparable to previously published studies [4, 5, 17, 18, 19]. The lack of an impact on non-COVID-19 mortality as a falsification test confirms the lack of bias when using this technique.

The main strength of our study is using an RDD approach to evaluate vaccine effectiveness. The RDD minimises the risk of bias due to unobserved factors driving both the assignment of treatment and the outcome. In this way, the fuzzy RDD is similar to a randomised control trial with imperfect compliance. The technique is particularly useful where it may be challenging to account for all confounding factors, such as in clinical records where the information on people’s socio-demographic characteristics are sparse. Studies investigating the effectiveness of the BCG vaccine against COVID-19 comparing COVID-19 outcomes in countries with different vaccine coverage rates, found a spurious association between vaccination coverage and COVID-19 mortality; an RDD approach based on birth cohort, which determined vaccine eligibility found no evidence of a protective effect, suggesting that unmeasured confounding factors drive the apparent effect on COVID-19 outcomes [10]. This is also a potential problem for COVID-19 vaccine effectiveness studies due to the differing characteristics, some unmeasured, between the vaccinated and unvaccinated populations. The RDD result therefore acts as a verification of observational studies and provides an unbiased estimate of the effectiveness. Another strength of our study is the use of population-wide data for England, covering approx. 93.9% of the population aged 75–84 years who received a first dose of a COVID-19 vaccine recorded in NIMS by 3 May 2021.

The main limitation of our study is that the RDD yielded an estimate of the vaccine effectiveness that is only valid for people aged around 80. It cannot be generalised to other age groups. Another limitation of the method is that people aged 75–79 years old rapidly become vaccinated, leaving a short window where the RDD can be applied. To mitigate the contamination of the control group, we estimated the discontinuity in mortality rates over a short time window (15 days), using dates where the difference in vaccination rates between people aged below and above 80 was the largest. We also estimated the discontinuity in weighted proportion of people who were vaccinated and were likely to have developed antibodies, rather than just focusing on the proportion of people who had received the vaccine. Nonetheless, our results may underestimate vaccine effectiveness. In addition, our analysis only provides an estimate of vaccine effectiveness for the first dose of COVID-19 vaccines. Another limitation is that our approach does not account for the different antibody responses in people who have previously been infected with COVID-19.

The RDD estimate of vaccine effectiveness against COVID-19 mortality of 52.6% is comparable to, but slightly lower than, previously reported by other studies based on different designs. In the UK, the effectiveness of a single dose of the Pfizer-BioNTech vaccine against COVID-19 mortality has been reported as 85% for 80+ year olds [17] and 80-85% for 70+ year olds, rising to 97% effective for two doses [5] [4]. The effectiveness of the Oxford-AstraZeneca vaccine has been reported as 80% in 70+ year olds [4]. International studies have reported effectiveness of 96.7% for the Pfizer-BioNTech vaccine after 7 days [18], and for mRMA vaccines 98.7% at least 7 days after the second dose, and 64.2% at least 14 days after one dose [20].

The RDD may underestimate vaccine effectiveness compared to methods investigating vaccine effectiveness on mortality with a delay after vaccination, due to deaths occurring soon after a person has been vaccinated caused by an infection prior to vaccination. The weighting reduces the effect of such deaths, but does not completely eliminate it, and we cannot determine when an infection has been acquired. In contrast, the published studies may over-estimate the vaccine effectiveness, by not accounting for all confounding factors.

Typically, RDD based studies of vaccine effectiveness have been applied where a policy change generated a cut-off in vaccine eligibility for a particular birth date, therefore outcomes such as mortality could be compared around this cut-off to determine their relationship to vaccination [10, 8]. The outcomes are measured on a much longer timescale than the time taken for the vaccine to produce protection in an individual and individuals do not change their vaccination status during the study. However, in this study, the rapid increase in vaccinations necessitated a short analysis period (here 15 days) and the changing status of individuals as they became vaccinated or developed higher levels of protection had to be taken into account. We used the proportion who were vaccinated for long enough to be likely to have reached threshold antibody level after one dose as an estimate of the proportion that are protected. However, there may be some level of protection for those who had not yet reached threshold level, and higher levels of protection for those who had developed greater than the threshold antibody level, possibly due to receiving a second dose. The number of second dose vaccinations in our data was small (less than 6% of people in our analysis dataset had received a second vaccination by the end of the analysis period), therefore, we only accounted for first vaccinations in the model.

Because COVID-19 vaccines are already approved and licensed for use, real-world vaccine effectiveness has to be estimated from observational data, which is challenging because of potential for residual confounding. The RDD estimates of vaccine effectiveness against COVID-19 mortality are unlikely to be biased by unmeasured confounding and are comparable to estimates obtained using different approaches. This suggests that residual confounding is unlikely to substantially bias estimates of vaccine effectiveness, at least for this age group.

## Data Availability

The ONS COVID-19 Public Health Linked Data Asset is available on the ONS Secure Research Service for
Accredited researchers. Researchers can apply for accreditation through the Research Accreditation
Service. The derived data used to estimate the RDD will be made publicly available.

## Acknowledgement

We thank Julie Stanborough, Emma Rourke and Ian Diamond for useful discussions about this work.

## Ethics approval

Ethical approval was obtained from the National Statistician’s Data Ethics Advisory Committee (NSDEC(20)12)

## Data sharing statement

The ONS COVID-19 Public Health Linked Data Asset is available on the ONS Secure Research Service for Accredited researchers. Researchers can apply for accreditation through the Research Accreditation Service. The derived data used to estimate the RDD will be made publicly available.

## Author contributors

VN led the study conceptualisation and design. JM led the preparation of the study data, in collaboration with CB. CB performed the statistical analyses, which were checked by VN. CB and VN drafted the manuscript. All authors contributed to interpretation of the results. All authors contributed to the critical revision of the manuscript. All authors approved the final manuscript. VN is the guarantor for the study. The corresponding author attests that all listed authors meet authorship criteria and that no others meeting the criteria have been omitted.

## Conflict of interest statement

None

## Footnotes

1 https://coronavirus.data.gov.uk/details/vaccinations

## References

[1] F. P. Polack, S. J. Thomas, N. Kitchin, J. Absalon, A. Gurtman, S. Lockhart, J. L. Perez, G. Pérez Marc, E. D. Moreira, C. Zerbini, R. Bailey, K. A. Swanson, S. Roychoudhury, K. Koury, P. Li, W. V. Kalina, D. Cooper, R. W. Frenck, L. L. Hammitt, Ö. Türeci, H. Nell, A. Schaefer, S. Ünal, D. B. Tresnan, S. Mather, P. R. Dormitzer, U. Şahin, K. U. Jansen and W. C. Gruber, “Safety and Efficacy of the BNT162b2 mRNA Covid-19 Vaccine,” New England Journal of Medicine, vol. 383, no. 27, pp. 2603-2615, 12 2020.

[2] M. Voysey, S. A. C. Clemens, S. A. Madhi, L. Y. Weckx, P. M. Folegatti and P. K. e. a. Aley, “Safety and efficacy of the ChAdOx1 nCoV-19 vaccine (AZD1222) against SARS-CoV-2: an interim analysis of four randomised controlled trials in Brazil, South Africa, and the UK,” The Lancet, vol. 397, no. 10269, pp. 99-111, 1 2021.

[3] L. R. Baden, H. M. El Sahly, B. Essink, K. Kotloff, S. Frey, R. Novak, D. Diemert, S. A. Spector, N. Rouphael, C. B. Creech, J. McGettigan, S. Khetan, N. Segall, J. Solis, A. Brosz, C. Fierro, H. Schwartz, K. Neuzil, L. Corey, P. Gilbert, H. Janes, D. Follmann, M. Marovich, J. Mascola, L. Polakowski, J. Ledgerwood, B. S. Graham, H. Bennett, R. Pajon, C. Knightly, B. Leav, W. Deng, H. Zhou, S. Han, M. Ivarsson, J. Miller and T. Zaks, “Efficacy and Safety of the mRNA-1273 SARS-CoV-2 Vaccine,” New England Journal of Medicine, vol. 384, no. 5, pp. 403-416, 2 2021.

[4] J. Lopez Bernal, N. Andrews, C. Gower, J. Stowe, E. Tessier, R. Simmons and Mary Ramsay, “Effectiveness of BNT162b2 mRNA vaccine and ChAdOx1 adenovirus vector vaccine on mortality following COVID-19,” Medrxiv, 2021.

[5] J. Lopez Bernal, N. Andrews, C. Gower, C. Robertson, J. Stowe, E. Tessier, R. Simmons, S. Cottrell, R. Roberts, M. O’Doherty, K. Brown, C. Cameron, D. Stockton, J. McMenamin and M. Ramsay, “Effectiveness of the Pfizer-BioNTech and Oxford-AstraZeneca vaccines on covid-19 related symptoms, hospital admissions, and mortality in older adults in England: test negative case-control study,” BMJ, p. n1088, 5 2021.

[6] E. Vasileiou, C. R. Simpson, T. Shi, S. Kerr, U. Agrawal, A. Akbari, S. Bedston, J. Beggs, D. Bradley, A. Chuter, S. de Lusignan, A. B. Docherty, D. Ford, F. R. Hobbs, M. Joy, S. V. Katikireddi, J. Marple, C. McCowan, D. McGagh, J. McMenamin, E. Moore, J. L. Murray, J. Pan, L. Ritchie, S. A. Shah, S. Stock, F. Torabi, R. S. Tsang, R. Wood, M. Woolhouse, C. Robertson and A. Sheikh, “Interim findings from first-dose mass COVID-19 vaccination roll-out and COVID-19 hospital admissions in Scotland: a national prospective cohort study,” 5 2021. [Online].

[7] N. Dagan, N. Barda, E. Kepten, O. Miron, S. Perchik, M. A. Katz, M. A. Hernán, M. Lipsitch, B. Reis and R. D. Balicer, “BNT162b2 mRNA Covid-19 Vaccine in a Nationwide Mass Vaccination Setting,” 4 2021. [Online].

[8] N. E. Basta and M. E. Halloran, “Evaluating the Effectiveness of Vaccines Using a Regression Discontinuity Design,” American Journal of Epidemiology, vol. 188, no. 6, pp. 987-990, 6 2019.

[9] J. A. Lopez Bernal, N. Andrews and G. Amirthalingam, “The Use of Quasi-experimental Designs for Vaccine Evaluation,” Clinical Infectious Diseases, vol. 68, no. 10, pp. 1769-1776, 5 2019.

[10] C. de Chaisemartin and L. de Chaisemartin, “Bacille Calmette-Guérin Vaccination in Infancy Does Not Protect Against Coronavirus Disease 2019 (COVID-19): Evidence From a Natural Experiment in Sweden,” Clinical Infectious Diseases, vol. 72, no. 10, pp. e501-e505, 5 2021.

[11] L. M. Smith, L. E. Lévesque, J. S. Kaufman and E. C. Strumpf, “Strategies for evaluating the assumptions of the regression discontinuity design: A case study using a human papillomavirus vaccination programme,” International Journal of Epidemiology, vol. 46, no. 3, pp. 939-949, 6 2017.

[12] WHO Headquarters (HQ), “Evaluation of COVID-19 vaccine effectiveness,” World Health Organization, 2021.

[13] Department of Health and Social Care, “Joint Committee on Vaccination and Immunisation: advice on priority groups for COVID-19 vaccination, 30 December 2020.,” 2020. [Online]. Available: www.gov.uk/government/publications/priority-groups-for-coronavirus-covid-19-vaccination-advice-from-the-jcvi-30-december-2020/joint-committee-on-vaccination-and-immunisation-advice-on-priority-groups-for-covid-19-vaccination-30-december-2020#contents.

[14] J. Wei, N. Stoesser, P. C. Matthews, R. Studley, I. Bell, J. I. Bell, J. N. Newton, F. Jeremy, I. Diamond, E. Rourke, A. Howarth, B. D. Marsden, S. Hoosdally, E. Y. Jones, D. I. Stuart, D. W. Crook, T. E. Peto, K. B. Pouwels, D. W. Eyre and A. S. Walker, “The impact of SARS-CoV-2 vaccines on antibody responses in the general population in the United Kingdom,” Nature Microbiology, vol. In press, 2021.

[15] D. S. Lee and T. Lemieux, “Regression Discontinuity designs in economics,” Journal of Economic Literature, vol. 48, no. 2, pp. 281-355, 6 2010.

[16] A. K. Clift, C. A. Coupland, R. H. Keogh, K. Diaz-Ordaz, E. Williamson, E. M. Harrison, A. Hayward, H. Hemingway, P. Horby, N. Mehta, J. Benger, K. Khunti, D. Spiegelhalter, A. Sheikh, J. Valabhji, R. A. Lyons, J. Robson, M. G. Semple, F. Kee, P. Johnson, S. Jebb, T. Williams and J. Hippisley-Cox, “Living risk prediction algorithm (QCOVID) for risk of hospital admission and mortality from coronavirus 19 in adults: national derivation and validation cohort study,” The BMJ, vol. 371, 10 2020.

[17] Public health England, “PHE COVID-19 vaccine effectiveness report March 2021,” 2021. [Online]. Available: https://assets.publishing.service.gov.uk/government/uploads/system/uploads/attachment_data/file/989360/PHE_COVID-19_vaccine_effectiveness_report_March_2021_v2.pdf.

[18] E. J. Haas, F. J. Angulo, J. M. McLaughlin, E. Anis, S. R. Singer, F. Khan, N. Brooks, M. Smaja, G. Mircus, K. Pan, J. Southern, D. L. Swerdlow, L. Jodar, Y. Levy and S. Alroy-Preis, “Impact and effectiveness of mRNA BNT162b2 vaccine against SARS-CoV-2 infections and COVID-19 cases, hospitalisations, and deaths following a nationwide vaccination campaign in Israel: an observational study using national surveillance data,” The Lancet, vol. 397, no. 10287, pp. 1819-1829, 5 2021.

[19] F. S. Vahidy, M. Mph, L. Pischel, M. E. Tano, A. P. Pan, M. L. Boom, H. D. Sostman, K. Nasir, M. Mph, S. B. Omer and A. Director, “Real World Effectiveness of COVID-19 mRNA Vaccines against Hospitalizations and Deaths in the United States,” Medrxiv, 2021.

[20] G. Chodick, L. Tene, T. Patalon, S. Gazit, A. Ben Tov, D. Cohen and K. Muhsen, “The Effectiveness of the first dose of BNT162b2 vaccine in reducing SARS-COV-2 infection 12-24 days after immunization: real world evidence,” JAMA Netw Open, vol. 4, no. 6, 2021.

