## Supplementary Information for "Estimating the effectiveness of first dose of COVID-19 vaccine against mortality in England: a quasi-experimental study"

### Supplementary Figures

**Supplementary Figure 1: Cumulative incidence of second dose COVID-19 vaccination by age group (75–79, 80–84 years) from 8 December 2020 (date of first UK vaccination) to 3 May 2021.**

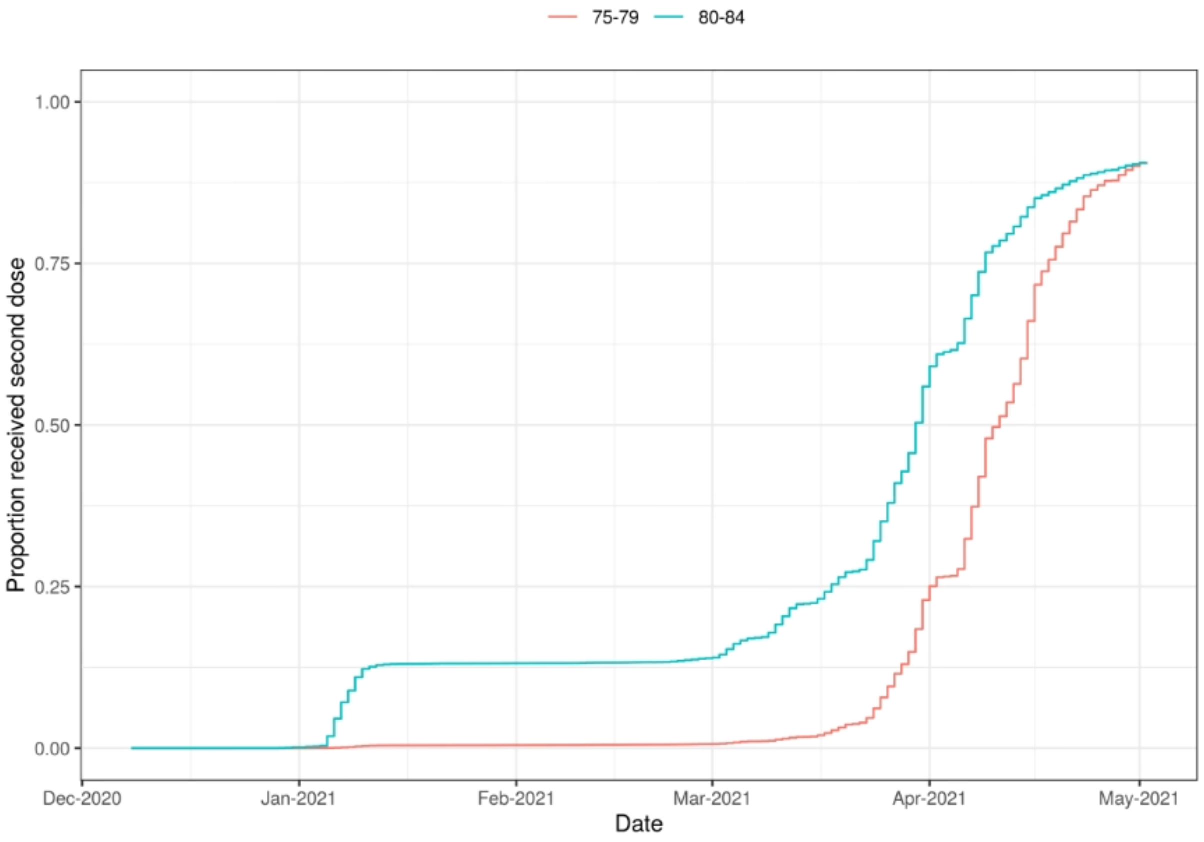


**Supplementary Appendix 1: Continuity of covariates**

An RDD analysis relies on there being no discontinuities in covariates at the eligibility cut off, as these could bias the results. To test this is correct for our analysis, the covariates sex, IMD quintile and whether someone is clinically extremely vulnerable are checked for continuity across the eligibility cut-off. The proportions of people in the dataset who are female, who live in the most deprived IMD quintile and who are clinically extremely vulnerable are plotted in Figure S2. Fitting linearly with a discontinuity at 80 years does not yield a statistically significant discontinuity for any of these variables.

**Supplementary Figure S2 Continuity across the eligibility cut-off for main covariates**

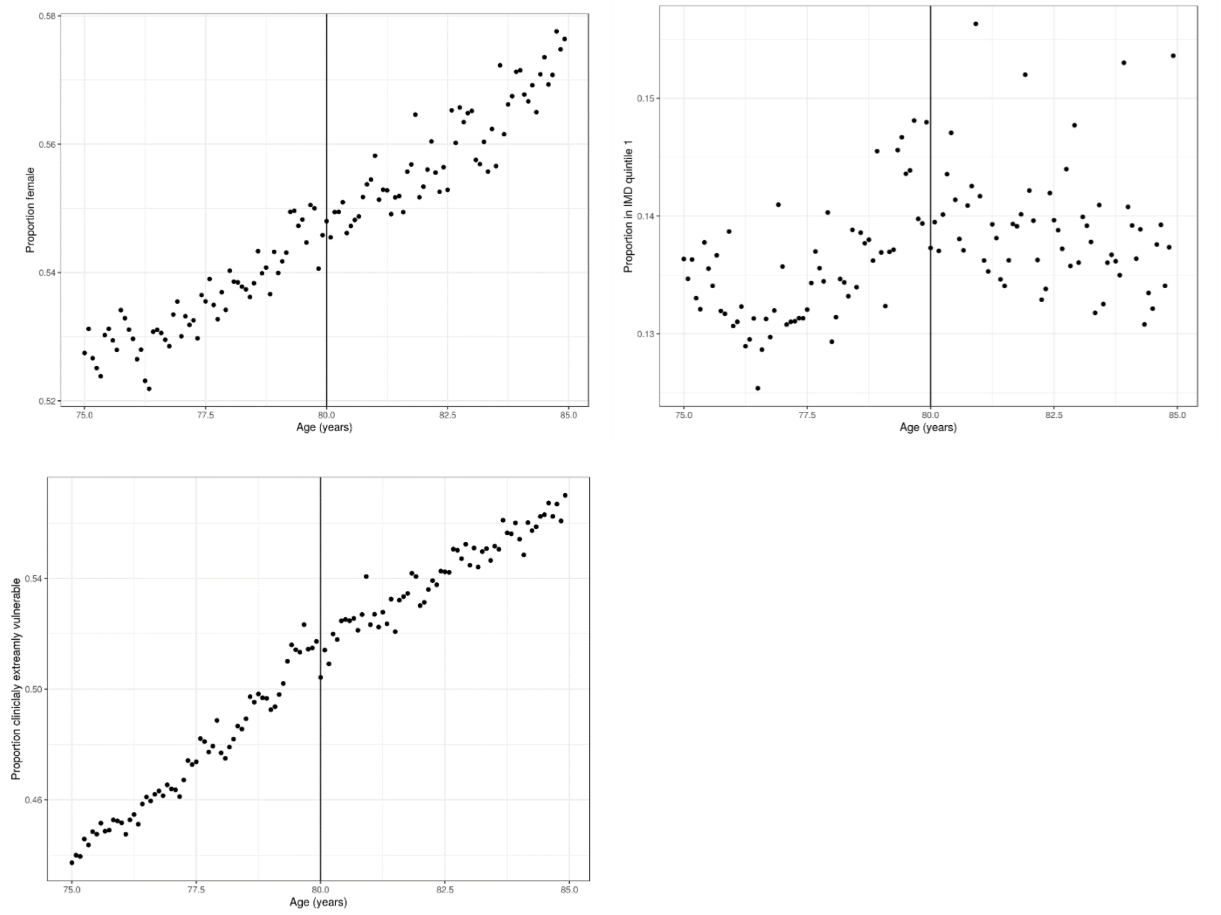


Figure S2: **A:** Proportion of females by age in months. **B:** Proportion of people in IMD quintile 1 by age in months. **C:** Proportion of people who are clinically extremely vulnerable by age in months. The p values for the discontinuity from the linear fit with a discontinuity at age 80 are 0.913, 0.743 and 0.826 respectively.

**Supplementary Appendix 2: Method for calculating the weighted proportion of people 'treated'**

Cumulative incidence curves are used to calculate the probability of having been vaccinated 0/7/14/21/28 days prior to each day of the analysis period for each age in months age group, for each vaccine manufacturer (Pfizer and AstraZeneca). People vaccinated before the analysis period are included on day zero. For each day in the analysis period, the proportions of people with a delay of 28 days post vaccination is weighted by the proportion of people who reach threshold antibody level for the corresponding number of days post vaccination using data from the CIS study for that vaccine [14]. The additional proportion of people who have been vaccinated for 21 days (compared to 28 days) is then weighted by the appropriate threshold antibody proportion and added to the weighted 28 days proportion. This is repeated for 21/14/7 and 0 days delay. This results in an estimated proportion of people who have reached threshold antibody level (hence are treated) on each day of the analysis period.

For days other than the first day of the analysis period, the estimated threshold antibody proportion is weighted again by the proportion of the analysis period that they are treated for. This results in an estimated proportion of people treated (reached threshold antibody level) for the analysis period.

Early Moderna vaccinations are not included in the analysis. Vaccinations with an unknown vaccine manufacturer are treated as Pfizer, as there are approximately 1.5 times more Pfizer vaccines than AstraZeneca in our analysis period dataset. These account for a very small percentage (0.01%) of the total vaccines administered the analysis period dataset.

The model was run with fixed definitions of 'treated' corresponding to reaching a certain number of days post vaccination, and weighting Pfizer and AstraZeneca vaccines equally. As the definition of treated includes more days post vaccination, the LATE and hence the vaccine effectiveness also increases. For low post vaccination delays, full protection is assumed soon after vaccination, however in reality this is unlikely to be the case so the mortality of people classed as treated but recently vaccinated will decrease the apparent vaccine effectiveness (Table S1). For long post vaccination delays, protection is assumed to only be present a long time after vaccination, whereas in reality it is likely to occur before, so the lower mortality of protected people at less than the post vaccination delay, who in reality have some protection, will contribute to a larger than expected vaccine effectiveness.

**Supplementary Table S1: Estimates of Local Average Treatment Effect and Vaccine effectiveness by post vaccination delay**

| Post vaccination delay | LATE /100,000 (95% CI) | VE (%) |
|---|---|---|
| 0 | -81 (-139, -23) | 41.7 (38.2, 89.0) |
| 7 | -67 (-115, -19) | 36.2 (44.9, 90.6) |
| 14 | -84 (-144, -25) | 43.0 (36.9, 88.0) |
| 21 | -105 (-178, -32) | 50.3 (29.0, 85.1) |
| 28 | -122 (-208, -37) | 55.8 (23.6, 82.7) |

**Supplementary Appendix 3: Effect of changing bandwidth and doughnut RDD**

The LATE for COVID-19 and non-COVID-19 mortality was calculated for different bandwidths around the eligibility cut-off of 960 months for the analysis period used in the main text. Bandwidths from 10 to 60 months were used. No bandwidth produced a statistically significant LATE for non-COVID mortality (Table S2). Bandwidths of 50 and 60 (as used in the main analysis) produced a statistically significant LATE for COVID-19 mortality with values of -112 per 100,000 (95% CI -202 – -21) and -122 per 100,000 (95% CI -191 – -33) respectively.

**Supplementary Table S2: Estimates of Local Average Treatment Effect on COVID-19 and non-COVID-19 mortality by bandwidths**

| | COVID-19 mortality | | Non COVID-19 mortality | |
|---|---|---|---|---|
| Bandwidth | LATE/100,000 deaths | p value | LATE/100,000 deaths | p value |
| 10 | -161 (-466 - 145) | 0.3 | 171 (-99 - 440) | 0.2 |
| 20 | -87 (-265 - 90) | 0.3 | 96 (-54 - 244) | 0.2 |
| 30 | -90 (-221 - 41) | 0.2 | 62 (-57 - 180) | 0.3 |

| | | | | |
|---|---|---|---|---|
| 40 | -85 (-191 - 21) | 0.1 | 79 (-23 - 182) | 0.1 |
| 50 | **-112 (-202 - -21)** | **0.02** | 40 (-57 - 105) | 0.4 |
| 60 | **-112 (-191 - -33)** | **0.005** | 24 (-57 - 105) | 0.6 |

A doughnut RDD with different widths of removed data was used to calculate the LATE for COVID-19 and non-COVID-19 mortality with a bandwidth of 60 around the age eligibility cut-off of 960 months. The effect of the size of the removed data does not significantly affect the LATE estimate, however the error on the LATE increases as more data is removed (Figure S3).

**Supplementary Figure S3: Estimates of Local Average Treatment Effect on COVID-19 removing data near cut-off for eligibility**

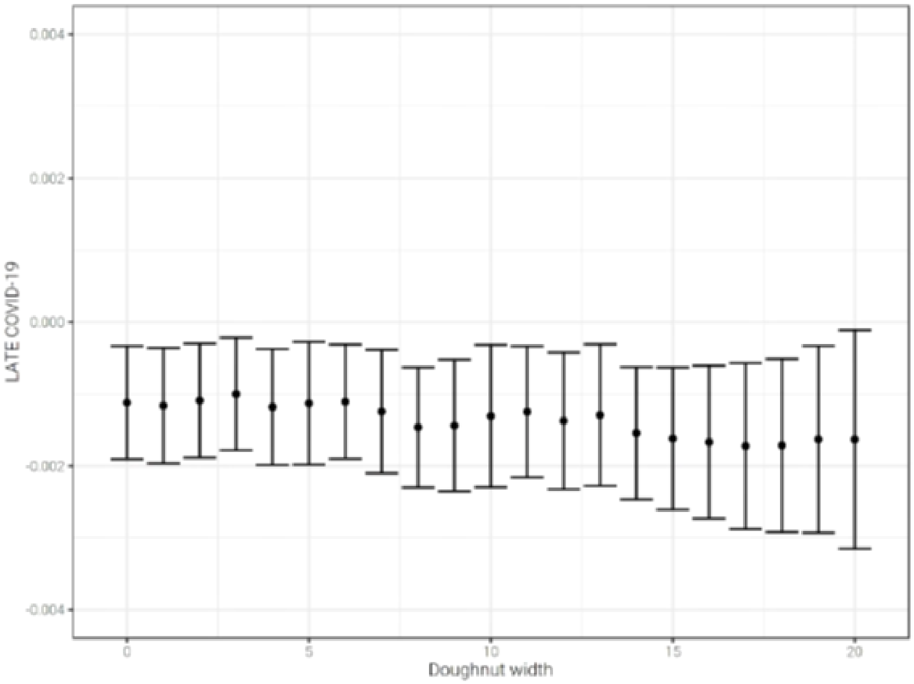


**Supplementary Appendix 4: Discontinuity in outcomes at other cut-offs**

COVID-19 mortality and non-COVID-19 mortality were tested for the presence of discontinuities at values of the instrumental variable other than the eligibility cut-off of 960 months for the analysis period used in the main text. RDDs were fit using cut-offs from 910 to 1010 months in 10-month intervals and the LATE found (Table S3). The LATE is not significant for non-COVID-19 mortality for any cut-off. The COVID-19 outcome data has a significant LATE only at age in months of 960, as we would expect due to the sharp discontinuity in the treatment probability at this cut-off.

**Supplementary Table S3: Estimates of Local Average Treatment Effect on COVID-19 and non-COVID-19 mortality for different cut-offs**

| | COVID-19 mortality | | Non COVID-19 mortality | |
|---|---|---|---|---|
| Cut-off (age in months) | LATE/100,000 | p value | LATE/100,000 | p value |
| 910 | 281 (-253, 814) | 0.3 | 205 (-402, 813) | 0.5 |

| | | | | |
|---|---|---|---|---|
| 920 | -142 (-385, 101) | 0.3 | -7 (-320, 306) | 1 |
| 930 | 172 (-189, 532) | 0.4 | -71 (-360, 502) | 0.7 |
| 940 | -701 (-2639, 1237) | 0.5 | -127 (-2077, 1824) | 0.9 |
| 950 | -152 (-322, 18) | 0.08 | 44 (-121, 210) | 0.6 |
| 960 | **-122 (-191, -33)** | **0.005** | 24 (-57, 105) | 0.6 |
| 970 | -135 (-323, 53) | 0.2 | -73 (-274, 127) | 0.5 |
| 980 | -716 (-3618, 2186) | 0.6 | 191 (-3804, 4186) | 0.9 |
| 990 | -555 (66, 12291) | 0.1 | 61 (-312, 435) | 0.7 |
| 1000 | -651 (228, 34586) | 0.3 | 83 (-295, 461) | 0.7 |
| 1010 | -660 (546, 85239) | 0.9 | -492, (-1099, 115) | 0.1 |

**Supplementary Appendix 5: Results with different length analysis periods**

Increasing the length of the analysis period has the effect of increasing the reliability of the COVID-19 mortality probability by age in months. However, due to the changing infection rate and the changing vaccination rate, it introduces more variation over the period for both the mortality and treatment measures. Therefore, the shortest analysis period that provides sufficient data for analysis of COVID-19 mortality is required.

Results for an analysis period start date of the 16 Jan 2021 (as for the analysis in the main text) and follow up times of 5, 10, 15, 20 and 25 days are shown in Table S4.

The relative uncertainty in the LATE is lowest for a 15 day follow up period. For shorter periods, there will not be enough mortality data to calculate a reliable estimate. For longer periods, the impact of the changing vaccination rate is greater.

**Supplementary Table S4: Estimates of Local Average Treatment Effect on COVID-19 mortality and Vaccine Effectiveness using different follow-up periods**

| Follow up (days) | LATE (/100,000) | Vaccine effectiveness (%) |
|---|---|---|
| 5 | -32 (-95, 30) | 55.9 (14.2, 136.5) |
| 10 | -66 (-133, -0.2) | 50.9 (25.3, 95.3) |
| 15 | -112 (-191, -33) | 52.6 (26.6, 84.3) |
| 20 | -95 (-178, -11) | 40.7 (36.0, 97.6) |
| 25 | -140 (-274, -6) | 47.5 (27.1, 102) |
